# Impaired Lymphocyte Responses in Pediatric Sepsis Vary by Pathogen Type

**DOI:** 10.1101/2021.09.15.21263652

**Authors:** Robert B. Lindell, Donglan Zhang, Jenny Bush, Douglas C. Wallace, Joshua D. Rabinowitz, Wenyun Lu, E. John Wherry, Scott L. Weiss, Sarah E. Henrickson

## Abstract

**Background:** Sepsis is the leading cause of death in hospitalized children worldwide. Despite its hypothesized immune-mediated mechanism, targeted immunotherapy for sepsis is not available for clinical use.

**Objective:** To determine the association between cytometric, proteomic, bioenergetic, and metabolomic abnormalities and pathogen type in pediatric sepsis.

**Methods:** Serial PBMC samples were obtained from 14 sepsis patients (34 samples) and 7 control patients for this pilot study. Flow cytometry was used to define immunophenotype, including T cell subset frequency and activation state, and assess intracellular cytokine production. Global immune dysfunction was assessed by TNF-α production capacity and monocyte HLA-DR expression. Mitochondrial function was assessed by bulk respirometry. Plasma cytokine levels were determined via Luminex assay. Metabolites were measured by liquid chromatography-mass spectrometry. Results were compared by timepoint and pathogen type.

**Results:** Sepsis patients were older and had higher illness severity compared to controls; demographics were otherwise similar. Compared to controls, sepsis patients demonstrated global immune dysfunction, loss of peripheral of non-naïve CD4^+^ T cells, and reduced PBMC mitochondrial function. Metabolomic findings in sepsis patients were most pronounced at sepsis onset and included elevated uridine and 2-dehydrogluconate and depleted citrulline. Loss of peripheral non-naïve CD4^+^ T cells was associated with immune dysfunction and reduced cytokine production despite increased T cell activation. CD4^+^ T cell differentiation and corresponding pro- and anti-inflammatory cytokines varied by pathogen.

**Conclusion:** Pediatric sepsis patients exhibit a complex, dynamic physiologic state characterized by immunometabolic dysregulation which varies by pathogen type.

## Introduction

Pediatric sepsis is the leading cause of death in hospitalized children worldwide.^1^ As in adults, sepsis in children is characterized by concurrent pro- and anti-inflammatory states with dysregulation of the innate and adaptive immune response to infection.^2,3^ Critical care for sepsis is limited to antibiotics, source control, and supportive care for organ dysfunction.^4^ Immunocompromised patients and those who develop immune suppression in the setting of sepsis both represent high-risk clinical phenotypes, with mortality rates that exceed 50%.^5,6^ Despite a hypothesized immune-mediated mechanism, successful interventional trials of targeted immunomodulation in sepsis remain elusive.^7^

Previous investigations have demonstrated that focused measures indicative of innate and adaptive immune dysfunction^8-10^ are associated with secondary infection,^11^ persistent organ dysfunction^12^ and mortality^5,6,13^ in pediatric sepsis, however comprehensive analyses of immunometabolic changes are lacking and could yield therapeutic insights for these high-risk patients. Advances in immune profiling have the potential to allow new insights into the role that lymphocytes play in shaping the immune response to sepsis.^14-16^ Here we report on a single-center prospective, observational study of immunometabolic function in children with sepsis in which we identified sepsis-associated immune dysregulation which varied by pathogen type through longitudinal cytometric, proteomic, bioenergetic, and metabolomic assays.

## Patients and Methods

### Study Design and Population

The study was approved by the Children’s Hospital of Philadelphia Institutional Review Board, and written informed consent was obtained prior to any study procedures. We performed a prospective observational study of patients less than 18 years treated for severe sepsis or septic shock in a single academic PICU between May 2014 and June 2018. Severe sepsis and septic shock were defined using consensus pediatric criteria.^2^ Exclusion criteria were weight less than 7.5kg (due to limits on blood collection), leukocyte count less than 0.5×10^3^/μL, known mitochondrial disease, unrepaired cyanotic heart disease, and prior study enrollment. A convenience sample of postoperative neurosurgical patients without evidence of infection or organ dysfunction were enrolled as controls.

### Clinical Data Collection

Clinical data related to patient characteristics, therapies, and vital status were abstracted from each study participant. Organ dysfunction, as previously defined, was monitored for 28 days after sepsis recognition. Severity of illness was assessed by the Pediatric Index of Mortality-2 (PIM-2) score,^17^ Pediatric Risk of Mortality-III (PRISM-III) score,^18^ and Pediatric Logistic Organ Dysfunction (PELOD) score.^19^ Exposure to endotracheal intubation and mechanical ventilation were obtained from clinical flowsheets and physician documentation. Exposure to vasopressor infusion and steroids were obtained from the medication administration record.

### Biospecimen Collection

A day 1-2 sample of 7-9ml of blood was collected as soon as possible after consent and enrollment. For patients with sepsis, additional blood was collected between study days 3-5 (and at least 2 days after first sample) and again between days 8-14 for patients who remained in the PICU. For control patients, a single sample was drawn on the date of consent and enrollment. Blood was collected in citrate tubes for measurement of mitochondrial respiration, EDTA tubes for monocyte human leukocyte antigen DR (HLA-DR) expression, and lithium heparin tubes (on ice) for measurement of whole-blood *ex vivo* LPS-induced tumor necrosis factor-alpha (TNF-α) and plasma cytokine analyses. Mitochondrial respiration, monocyte HLA-DR, and *ex vivo* LPS-stimulated TNF-α were measured on fresh blood samples. Additional peripheral blood mononuclear cells (PBMCs) and heparin plasma were isolated by density gradient centrifugation and cryopreserved at -80 degrees C for subsequent analysis.

### Flow Cytometry

Cryopreserved PBMCs were partially thawed in a 37°C water bath, then resuspended in warm complete medium and plated into a 96-well round-bottom plate at a concentration of 1×10^6^ PBMCs per well. For general immune profiling, cells were incubated with a cocktail of antibodies targeting surface proteins at room temperature for 30 minutes and then washed. After fixation and permeabilization, cells were incubated with a cocktail of antibodies targeting intracellular proteins at room temperature for 60 minutes and then washed. After staining, cells were resuspended into 1.6% PFA fixative and held overnight at 4°C prior to acquisition. For analysis of intracellular cytokines, PBMCs were stimulated with PMA/ionomycin for 4 hours prior to surface and intracellular staining per the above protocol.

Cells were analyzed on a LSR II flow cytometer (BD Biosciences, San Jose, CA) according to the manufacturer protocol. Single stain controls were performed with compensation beads (UltraComp Beads, Invitrogen/ThermoFisher Scientific, Waltham, MA). Compensated FCS files were then loaded into FlowJo (Tree Star, Ashland, OR) for predefined analysis of immune cell subsets. After identification of live singlet CD3+ lymphocytes, populations of CD4^+^ and CD8^+^ lymphocytes were identified through bivariate plots. Subsets of CD4+ and CD8+ cells were analyzed by CD45RA and CD27 expression using the following definitions: naïve T cell (CD45RA^+^/CD27^+^), central memory T cell (CD45RA^-^/CD27^+^), effector memory T cell (CD45RA^-^/CD27^-^), and TEMRA cell (CD45RA^+^/CD27^-^) which are a subset of effector memory cells which re-express CD45RA. Non-naïve CD4^+^ and CD8^+^ T cells were defined as the union of central memory T cells, effector memory T cells, and TEMRA cells.

We then assessed CD4+ T cell differentiation by measuring intracellular cytokines via flow cytometry after PMA/ionomycin stimulation. After defining non-naïve T cells using the lineage markers and gating strategy above, we identified CD4^+^ T cell states based on intracellular cytokine expression: Th1 (IFN-γ^+^), Th2 (IL13^+^), Th17 (IL-17α^+^), Tfh (IL-21^+^), and Trseg (FoxP3^+^). Finally, in this group we defined the activation state of these CD4+ T cell subsets by defining the IL-2^+^ and CD38^+^ cells within groups.

### Mitochondrial Bioenergetics

Mitochondrial respiration was measured in PBMCs isolated from citrated whole blood by density gradient centrifugation as previously described.^20^ PBMC cell counts were performed using trypan blue exclusion (Countess II, Life Technologies, Grand Island, NY) with median viability 88% (interquartile range 76-95%). After isolation, the PBMC pellet was re-suspended in Hank’s balanced salt solution (pH 7.40) containing 5.5 mM glucose, 1mM pyruvate, and 10 mM HEPES, centrifuged a final time at 100g for 10 minutes at 20°C, and then again re-suspended in the same “respiration buffer”. Mitochondrial respiration was measured in 2-4×10^6^ intact PBMCs at 37°C using a high-resolution oxygraph (Oxygraph-2k Oroboros Instruments, Innsbruck, Austria). Oxygen flux (in pmol O_2_/sec/10^6^ cells), which is directly proportional to oxygen consumption (respiration), was recorded continuously using DatLab software 4.3 (Oroborus Instruments, Innsbruck, Austria) as shown below and as previously described.^20,21^ Intact cells were utilized to maintain the cellular microenvironment such that respiration relied on endogenous substrates. After routine oxygen consumption was recorded for 10-20 minutes, the ATP-synthase inhibitor oligomycin (1 μg/mL) was added to induce a state 4-like respiration independent of mitochondrial ATP production. Under these conditions, respiration was primarily due to leakage of protons across the inner mitochondrial membrane (LEAK). Routine minus LEAK indicated ATP-linked respiration. Maximal oxygen consumption through the electron transport system (ETS) was obtained by stepwise titration (1-2 μM) of the uncoupler carbonyl cyanide m-chlorophenylhydrazone (CCCP) until no further increase in oxygen consumption was detected (ETS_max_). The ETS_max_ indicates the maximal oxygen consumption possible through the electron transport system after pharmacologically uncoupling oxygen utilization from ATP production to assess the integrity of the ETS independent of energy production. Therefore, ETS_max_ reflects only the ability the ETS to use electrons to reduce oxygen to water without the need to add high-energy phosphate bonds to produce ATP, a final step in the energy-production pathway that normally helps to regulate ETS activity. Mitochondrial respiration was then inhibited by adding the ETS complex IV inhibitor, sodium azide, in 5-10 mM increments, followed by the ETS complex III inhibitor, antimycin A, in 5μL increments until no further decrease in oxygen consumption was observed. The residual oxygen flux reflective of non-mitochondrial respiration was subtracted from other respiration values. Respiration supporting mitochondrial ATP synthesis (ATP-linked respiration) was calculated as routine minus LEAK respiration. Spare respiratory capacity (SRC), calculated as ETS_max_ minus routine respiration, is the mitochondrial bioenergetic reserve available for cells to produce ATP in response to a stress-induced increase in metabolic demand.

### Functional Assays

*Ex vivo* LPS-stimulated whole blood TNF-α was measured by mixing 50ml heparinized whole blood with 500ml (250 pg) of phenol-extracted LPS from *Salmonella enterica abortus equi* (Sigma-Aldrich, L5886) within 60 to 90 minutes of blood collection as previously described ^8^. The sample was then incubated for 4 hours at 37°C, followed by centrifugation at 1,000 g for 5 minutes. The supernatant was stored at -80°C for batched analysis. TNF-α was measured, in duplicate, using an enzyme-linked immunosorbent assay kit (Invitrogen KHC3011C). Immunoparalysis was *a priori* defined as LPS-stimulated TNF-α less than 200pg/ml. Monocyte HLA-DR was measured using a whole-blood lysis technique. The sample was then stained with labeled anti-HLA-DR and anti-CD14 antibodies and the percentage of HLA-DR-positive cells among the CD-14 positive population was determined using flow cytometry.

### Plasma Proteomics

Interleukin (IL)-4, IL-6, IL-10, IL-18, IL-1RA, and Interferon (IFN)-γ were measured using commercially available assays (Cytokine Human Magnetic 30-Plex Panel, Invitrogen/ThermoFisher Scientific, Waltham, MA; Human Magnetic Luminex Assay and CRP DuoSet ELISA, R&D Systems, Minneapolis, MN). All samples were run in duplicate. FLUOstar software® (Ortenberg, Germany) was used to calculate standard curves for each analyte. To ensure accuracy and precision, the standard curve was required to have r^2^ >95%, and the results from mock samples were required to be within the 95% confidence interval of the expected range provided by the manufacturer.

### Plasma Metabolomics

Water-soluble plasma metabolites were analyzed by liquid chromatography-mass spectrometry, using the protocol previously described by Lu et al.^22^ Briefly, 200ml ice-cold methanol was added to each 50μl plasma sample, vortexed for 10 seconds, and then transferred to a -20°C for 20 minutes. Samples were centrifuged at 13,200rpm for 15 minutes. The supernatant was extracted and then dried under nitrogen flow and reconstituted in 1ml LC-MS grade water. Samples were analyzed on two separate instrument platforms to cover both positive charged and negative charged metabolites.

Negative charged metabolites were analyzed via reverse-phase ion-pairing chromatography coupled to an Exactive orbitrap mass spectrometer (ThermoFisher Scientific, San Jose, CA). The mass spectrometer was operated in negative ion mode with resolving power of 100,000 at m/z 200, scanning range being m/z 75-1000. The LC separation was achieved using a Synergy Hydro-RP column (100mm × 2 mm, 2.5 µm particle size, Phenomenex, Torrance, CA) with a flow rate of 200 µL/min. The LC gradient was 0 min, 0% B; 2.5 min, 0% B; 5 min, 20% B; 7.5 min, 20% B; 13 min, 55% B; 15.5 min, 95% B; 18.5 min, 95% B; 19 min, 0% B; 25 min, 0% B. Solvent A is 97:3 water:methanol with 10 mM tributylamine and 15 mM acetic acid; solvent B is methanol.

Positive charged metabolites were analyzed on a Q Exactive Plus mass spectrometer coupled to Vanquish UHPLC system (ThermoFisher Scientific, San Jose, CA). The mass spectrometer was operated in positive ion mode with resolving power of 140,000 at m/z 200, scanning range being m/z 75-1000. The LC separation was achieved on an Agilent Poroshell 120 Bonus-RP column (150 × 2.1 mm, 2.7 µm particle size). The gradient was 0 min, 50 µL/min, 0.0%B; 6 min, 50 µL/min, 0% B; 12 min, 200 µL/min, 70% B; 14 min, 200 µL/min, 100 %B; 18 min, 200 µL/min, 100% B; 19 min, 200 µL/min, 0% B; 24 min, 200 µL/min, 0% B; 25 min, 50 µL/min, 0% B. Solvent A is 10mM ammonium acetate + 0.1% acetic acid in 98:2 water:acetonitrile and solvent B is acetonitrile.

For each sample, ion-specific chromatograms for metabolites of interest were identified, and ion counts were abstracted for further analysis. The following metabolites were analyzed in this exploratory analysis: (iso)leucine, alanine, arginine, aspartate, citrate, citrulline, glucosamine, glutamine, histidine, hypoxanthine, lactate, lysine, methionine, phenylalanine, proline, taurine, threonine, valine, 2-dehydrogluconate, 2-hydroxy-glutarate, acetyl-glycine, a-ketoglutarate, allantoin, asparagine, cystathionine, cysteine, D-glucarate, D-gluconate, fumarate, gluconolactone, glutamate, glycine, glycolate, hippuric acid, homocysteine/ methylcysteine, hydroxyphenylpyruvate, hydroxyproline, indolelactic acid, inosine, kynurenine, malate, N-acetyl-L-alanine, N-acetyl-L-ornithine, phenyllactic acid, pyruvate, serine, trehalose, tyrosine, uric acid, uridine, and xanthine.

### Statistical Analyses

Manual flow cytometry gating was completed in FlowJo^23^ as detailed above. Data processing and analyses were completed in R Studio.^24^ Extraction and computation was facilitated with the ‘dplyr’ package. Graphs were plotted with the ‘ggplot2’ and ‘ggpubr’ packages. All remaining source code was custom developed. Data are presented as median with interquartile range (IQR) for continuous variables and counts with percentages for categorical variables. Correlation coefficients were quantified by the Spearman rank correlation coefficient. Tests of association between continuous versus categorical variables were performed by unpaired Wilcoxon test or Kruskal-Wallis test as appropriate. Association between categorical variables was assessed by Fisher’s exact test. All tests were performed in a two-sided manner, using a nominal significance threshold of *p*<0.05 unless otherwise specified, with Bonferroni corrections for analyses that included multiple comparisons.

## Results and Discussion

### Study patients

Fourteen patients with septic shock and seven non-infected controls were included in our study. Serial blood samples were collected from patients with sepsis and processed for peripheral blood mononuclear cell (PBMC) and plasma; in total, 34 sepsis samples and 7 control samples were obtained. Sepsis samples were collected on days 1-2, 3-5, and 8-14 after sepsis recognition. Control samples were collected from neurosurgical patients in the pediatric intensive care unit (PICU) without infection or organ dysfunction. Detailed descriptions of patient recruitment have been reported previously.^25,26^

Patient demographics, clinical characteristics, and laboratory values are summarized in Table 1. As expected, sepsis patients had higher illness severity at admission compared to controls as assessed by the Pediatric Index of Mortality-2 (PIM-2) score,^17^ Pediatric Risk of Mortality-III (PRISM-III) score,^18^ and Pediatric Logistic Organ Dysfunction (PELOD) score.^19^ Sepsis patients were also older than controls, but other demographics were similar. Sepsis patients more frequently received endotracheal intubation, vasoactive infusion, and corticosteroids. There was one mortality in the sepsis cohort, none among controls.

**Table 1.**
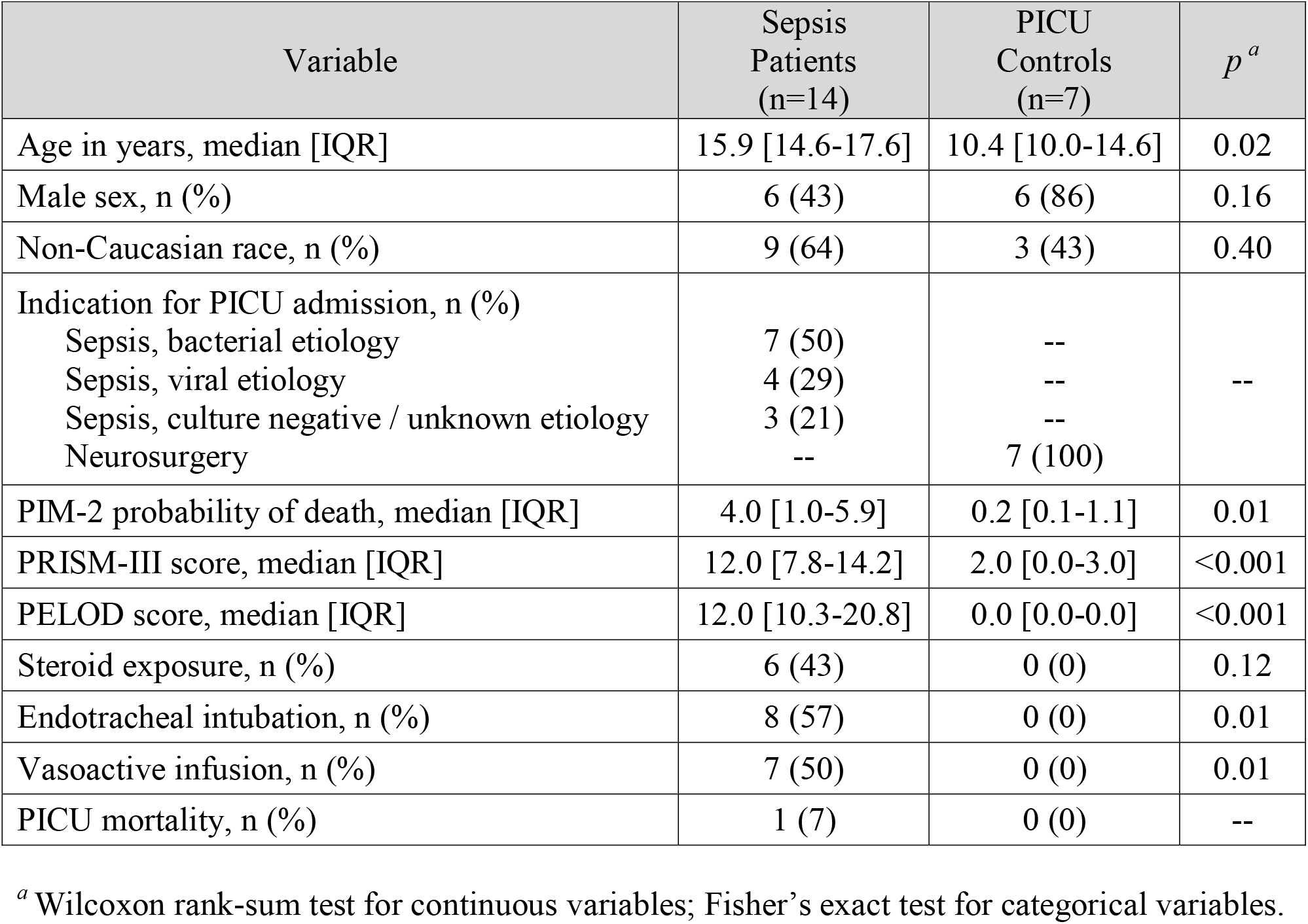
Characteristics of sepsis patients and controls at PICU admission

| Variable | Sepsis Patients<br>(n=14) | PICU Controls<br>(n=7) | *p*<sup>a</sup> |
| --- | --- | --- | --- |
| Age in years, median [IQR] | 15.9 [14.6-17.6] | 10.4 [10.0-14.6] | 0.02 |
| Male sex, n (%) | 6 (43) | 6 (86) | 0.16 |
| Non-Caucasian race, n (%) | 9 (64) | 3 (43) | 0.40 |
| Indication for PICU admission, n (%) |  |  |  |
| Sepsis, bacterial etiology | 7 (50) | -- | -- |
| Sepsis, viral etiology | 4 (29) | -- |  |
| Sepsis, culture negative / unknown etiology | 3 (21) | -- |  |
| Neurosurgery | -- | 7 (100) |  |
| PIM-2 probability of death, median [IQR] | 4.0 [1.0-5.9] | 0.2 [0.1-1.1] | 0.01 |
| PRISM-III score, median [IQR] | 12.0 [7.8-14.2] | 2.0 [0.0-3.0] | <0.001 |
| PELOD score, median [IQR] | 12.0 [10.3-20.8] | 0.0 [0.0-0.0] | <0.001 |
| Steroid exposure, n (%) | 6 (43) | 0 (0) | 0.12 |
| Endotracheal intubation, n (%) | 8 (57) | 0 (0) | 0.01 |
| Vasoactive infusion, n (%) | 7 (50) | 0 (0) | 0.01 |
| PICU mortality, n (%) | 1 (7) | 0 (0) | -- |

<sup>a</sup> Wilcoxon rank-sum test for continuous variables; Fisher's exact test for categorical variables.

### Pediatric sepsis patients exhibit global immunometabolic dysregulation

To quantify differences in lymphocyte subsets between sepsis patients and controls, we first assessed the abundance of key T cell subsets by flow cytometry (Fig. 1a). Compared to controls, the abundance of non-naïve CD4^+^ T cells was significantly reduced in patients with bacterial sepsis (Fig. 1b). Loss of central memory CD4^+^ T cells (CD45RA^-^/CD27^+^) was the primary driver of this finding; CD4^+^ TEMRA cells (CD45RA^+^/CD27^-^) were increased in patients with sepsis regardless of pathogen type. Non-naïve CD8^+^ T cell subsets were similar between sepsis and controls.

**Figure 1.**
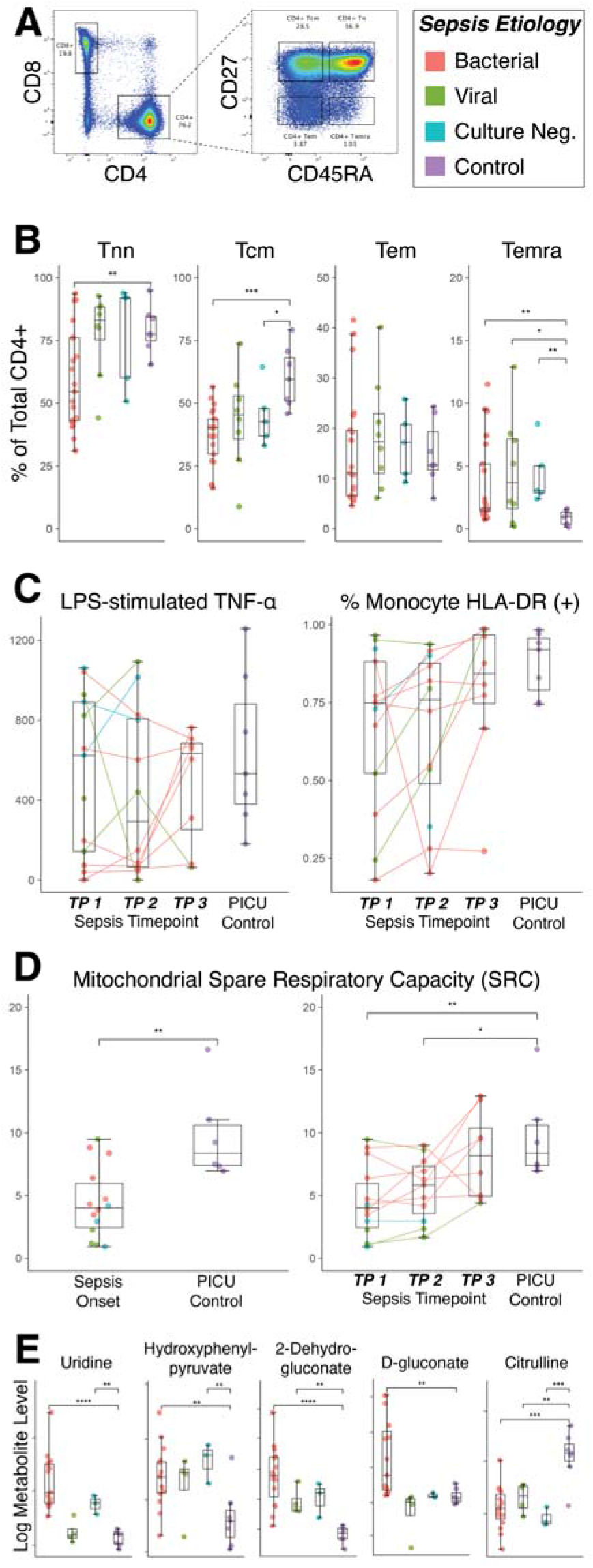
Immunometabolic dysregulation in pediatric sepsis patients. **Panel A:** CD4^+^ T cell gating strategy. **Panel B:** Relative abundance of T cell subsets differ between sepsis and control and are most pronounced for patients with bacterial sepsis. **Panel C:** Immune dysfunction at sepsis onset resolves in some patients on longitudinal sampling. **Panel D:** Mitochondrial function in sepsis patients improves through time. **Panel E:** Metabolomics findings reflect increased reactive oxygen species, amino acid breakdown products, and citrulline deficiency.

We then assessed two global markers of immune dysfunction commonly used in critical care – *ex vivo* lipopolysaccharide (LPS)-stimulated tumor necrosis factor (TNF)-α production capacity and monocyte human leukocyte antigen (HLA)-DR expression. Immunoparalysis, defined by TNF-α production capacity <200pg/ml,^8^ was more common in sepsis patients than controls (39% vs 0%, p=0.08). Median monocyte HLA-DR was lower at sepsis onset compared to controls (75% vs 92%, p=0.02). Cohort heterogeneity by interquartile range decreased on serial samples, and markers of immune dysfunction generally improved with sepsis recovery (Fig. 1c).

We interrogated bulk PBMC mitochondrial function using an Oroboros Oxygraph to measure spare respiratory capacity (SRC), which represents the mitochondrial bioenergetic reserve available for cells to produce ATP in response to stress-induced increase in metabolic demand.^27^ Median SRC was lower at sepsis onset compared to controls (4.0 vs 8.4, p=0.01). SRC improved through time and varied by pathogen type (Fig. 1d). Because the proportion of lymphocyte subsets varied between sepsis patients and controls, we constructed a generalized linear model which found no association between the relative proportion of CD4+ and CD8+ subsets (naïve, central memory, effector memory, TEMRA) in the PBMC sample and mitochondrial SRC, suggesting that differences in SRC are not confounded by shifts in cell subtypes.

Finally, we assessed plasma metabolites by liquid chromatography-mass spectrometry (Fig. 1e). In this exploratory analysis, some amino acid breakdown products including uridine (*p*<0.001) and hydroxyphenylpyruvate (*p*=0.012) and intermediate markers of glucose metabolism including 2-dehydrogluconate (*p*<0.001) and D-gluconate (*p*=0.017) were markedly elevated in patients with sepsis across timepoints compared to controls. Conversely, citrulline was decreased in children with sepsis regardless of pathogen (*p*<0.001); citrulline deficiency has been previously associated with impaired small bowel microcirculation, nitric oxide production, and mortality in sepsis.^28,29^

Taken together, these data demonstrate that pediatric patients with sepsis develop global immunometabolic dysregulation assessed by lower TNF-α production capacity, monocyte HLA-DR expression, mitochondrial SRC, loss of peripheral non-naïve T cells, and metabolomic abnormalities.

### T cell abundance and polarization in pediatric sepsis vary by pathogen type

Informed by the heterogeneity of immune abnormalities above, we then stratified T cell responses by pathogen type. Demographics and severity of illness did not vary by pathogen type. We assessed CD4^+^ T cell differentiation by measuring intracellular cytokines via flow cytometry after phorbol myristate acetate (PMA)/ionomycin stimulation and identified CD4^+^ T cell states based on resulting intracellular cytokine expression: Th1 (IFN-γ^+^), Th2 (IL13^+^), Th17 (IL-17α^+^), Tfh (IL-21^+^), Treg (FoxP3^+^). Th1 cells were reduced in bacterial sepsis and Th2 cells were reduced in viral sepsis compared to controls; Th17, Tfh, and Treg populations did not vary significantly (Fig. 2a). Th1 and Th2 lymphocyte abundance varied widely across serial samples (Fig. 2b). Among patients with bacterial sepsis, the proportion of IL2^+^ non-naïve CD4^+^ lymphocytes was lower at sepsis onset compared to controls (Fig. 2c), while the proportion of CD38^+^ cells did not vary by pathogen type.

**Figure 2.**
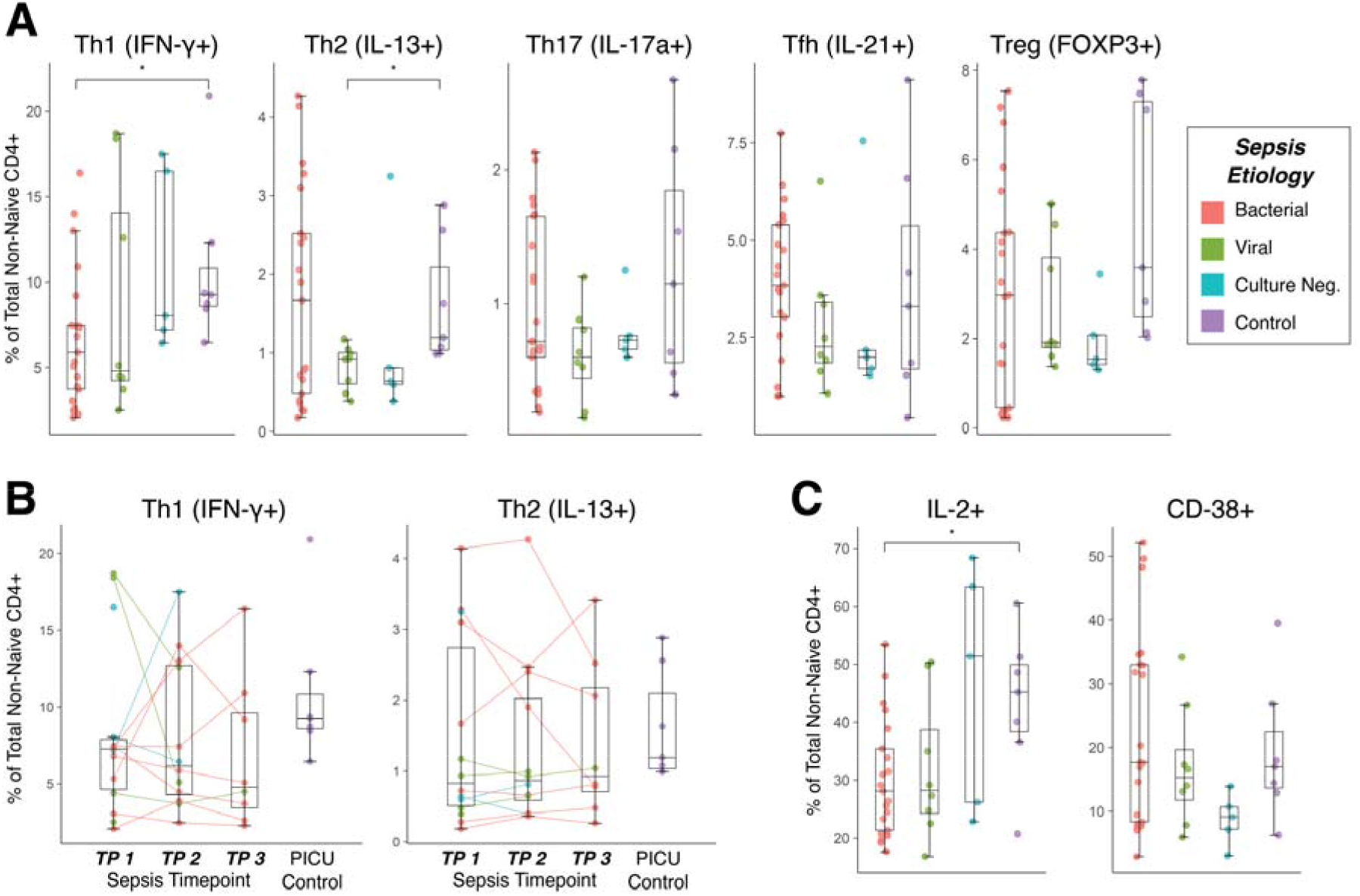
T cell activation and differentiation varies by pathogen type in pediatric sepsis patients. Panel. **A:** Differentiation of CD4^+^ T cells exhibits substantial heterogeneity which is partially explained by pathogen type. **Panel B:** In longitudinal sampling, Th1 and Th2 lymphocyte abundance varied within some patients but was constant in others. **Panel C:** Patients with bacterial sepsis had fewer IL-2+ lymphocytes than controls.

Both pro- and anti-inflammatory plasma cytokines were elevated at sepsis onset compared to controls and varied by pathogen type (Fig. 3). IL-6 and IL-18 were most elevated in bacterial sepsis; IFN-γ was low in this subgroup, consistent with reduced CD4+ Th1 cells in this population. IL-10 and IL-1RA were elevated in all sepsis patients, though IL-4 was only elevated in patients with viral sepsis. Among sepsis patients, cytokine values normalized through time as organ dysfunction resolved.

**Figure 3.**
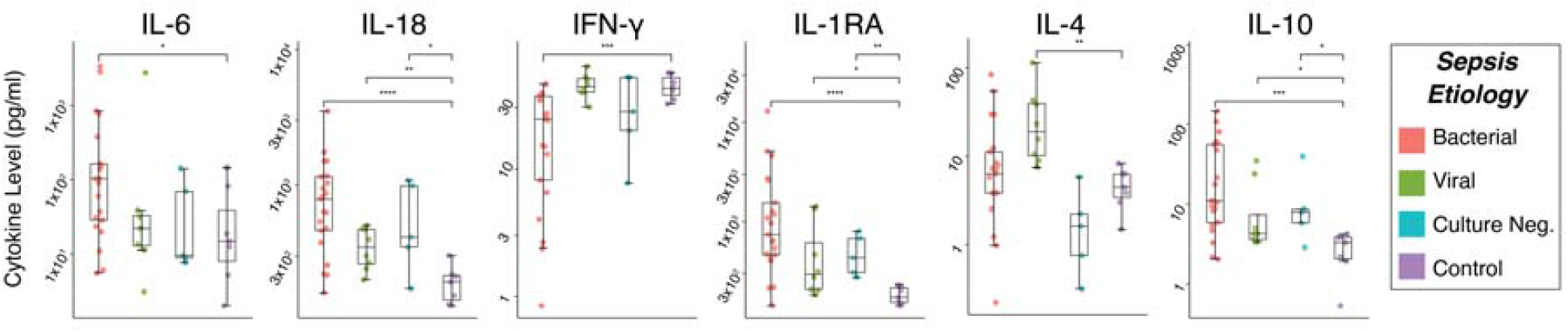
Plasma cytokine levels vary by pathogen type in pediatric sepsis patients.

### Loss of peripheral non-naïve CD4^+^ T cells is associated with immune dysfunction and characteristic immunometabolic dysregulation

In our final analysis, we examined the association between the loss of peripheral non-naïve CD4^+^ T cells and markers of immune dysfunction in sepsis patients. The loss of peripheral non-naïve CD4^+^ T cells is associated with both TNF-α production capacity (*R*=0.35, *p*=0.03) and monocyte HLA-DR expression (*R*=0.59, *p*<0.001; Fig. 4a). The frequency non-naïve CD4^+^ T cells is positively associated with serum IFN-γ level (*R*=0.57, *p*<0.001) and negatively associated with CD38 expression on non-naïve CD4^+^ T cells (*R*=-0.48, *p*=0.001), suggesting that loss of peripheral non-naïve CD4^+^ T cells is associated with decreased T cell function despite evidence of T cell activation (Fig. 4b).

**Figure 4.**
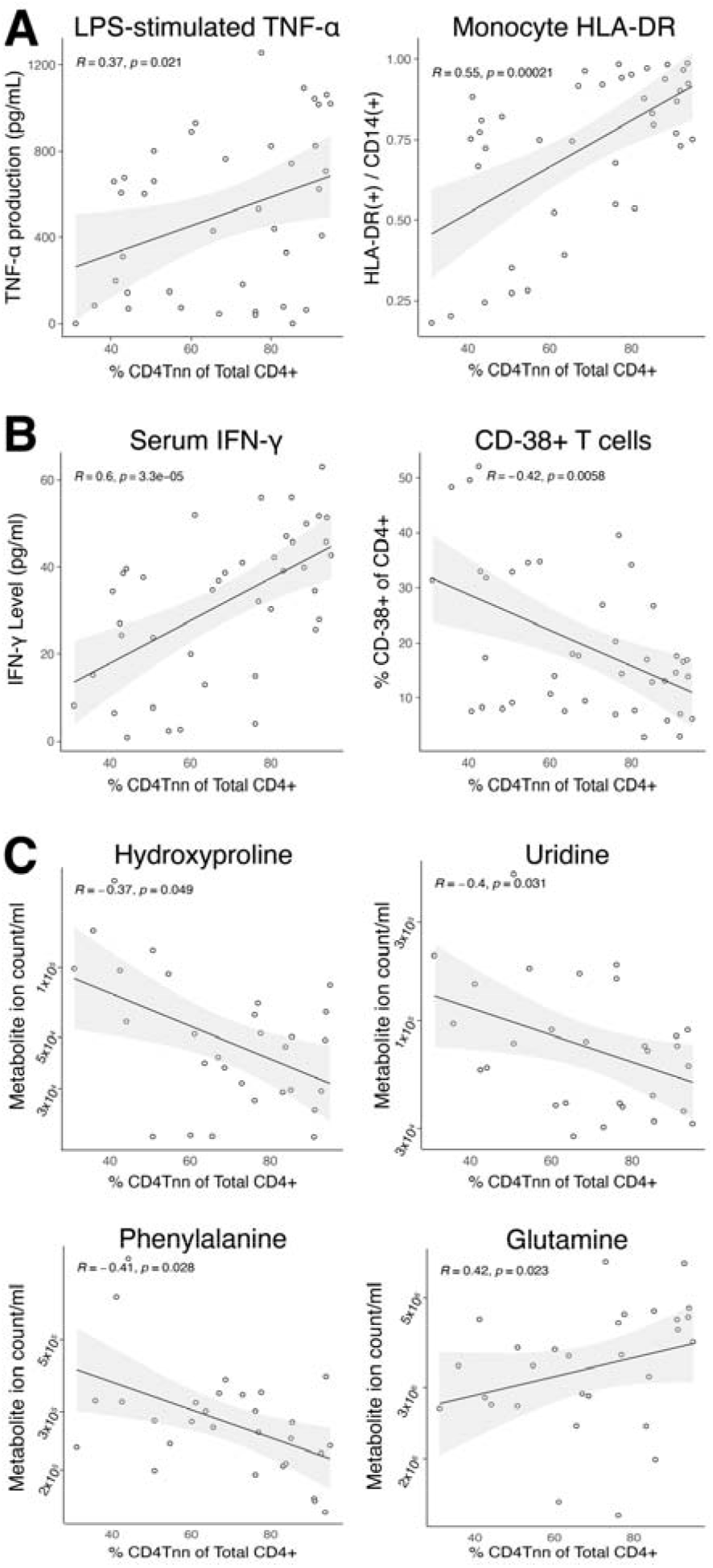
Loss of peripheral non-naïve CD4^+^ T cells is associated with functional, proteomic, and metabolomic abnormalities in pediatric sepsis patients. **Panel A:** The loss of peripheral non-naïve CD4^+^ T cells is associated with both TNF-production capacity and monocyte HLA-DR expression. **Panel B:** Non-naïve CD4^+^ T cells are positively associated with serum IFN-level and negatively associated with CD38 expression, suggesting that loss of peripheral non-naïve CD4^+^ T cells is associated with decreased T cell function. **Panel C:** Loss of peripheral non-naïve CD4^+^ lymphocytes is associated with several markers of protein catabolism and low plasma glutamine levels.

Finally, we tested the association between plasma metabolites and peripheral non-naïve CD4^+^ lymphocytes. The loss of peripheral non-naïve CD4^+^ T cells is associated with markers of protein catabolism (hydroxyproline, uridine, phenylalanine; Fig. 4c). Conversely, low glutamine was associated with decreased peripheral non-naïve CD4^+^ T cells. Glutamine is an essential cofactor for T cell activation, and glutamine depletion has been associated with reduced lymphocyte proliferation, cytokine production, and lymphocyte apoptosis.^30,31^

## Conclusions

We have demonstrated that pediatric sepsis is associated with characteristic immunometabolic dysregulation which vary by pathogen. Adaptive immune dysfunction in sepsis develops during a hyperinflammatory, catabolic state which is characterized by mitochondrial dysfunction and loss of peripheral non-naïve T cells. Heterogeneity in the immune response is partially explained by pathogen type, which appears to influence non-naïve T cell differentiation and function. These alternations in CD4^+^ T cell subset frequencies which vary by pathogen suggest that the dysregulated adaptive immune response to sepsis may be pathogen-specific.

While these findings are compelling, there are several limitations to our analysis of this pilot data. Our limited number of samples and cohort heterogeneity is susceptible to type II error. Comparison of sepsis patients to controls may bias toward the null if post-surgical patients have a mild inflammatory phenotype which overlaps with the immunometabolic phenotype of sepsis patients. Because steroid exposure was common in sepsis patients regardless of pathogen type, we cannot control for this important covariate in the present study. Because metabolic and mitochondrial measurements were performed in bulk, we cannot conclusively identify the association between these findings and specific immune cell subsets. Finally, we are unable to account for repeated measures within individuals due to limited sample size.

Through this pilot study, we have established the clinical feasibility of monitoring immune health in pediatric sepsis patients using small volume samples. Immune monitoring in pediatric critical illness is currently limited to inflammatory biomarkers and cytokine analysis. Deep immune profiling and functional testing of patient samples has the potential to identify mechanisms of immune dysfunction in pediatric sepsis, paving the way for personalized immunotherapy in critically ill children.

## Data Availability

Due to the nature of this research, participants of this study did not agree for their data to be shared publicly, so supporting data is not available.

## ABBREVIATIONS

PBMC: Peripheral blood mononuclear cells
PIM-2: Pediatric Index of Mortality-2
PRISM-III: Pediatric Risk of Mortality-III
PELOD: Pediatric Logistic Organ Dysfunction
LPS: Lipopolysaccharide
HLA-DR: Human leukocyte antigen - DR isotype
SRC: Spare respiratory capacity
PMA: Phorbol myristate acetate
Th1 cell: Type 1 CD4^+^ T helper cell
Th2 cell: Type 1 CD4^+^ T helper cell
Th17 cell: Type 1 CD4^+^ T helper cell
Tfh cell: T follicular helper cell
Treg cell: Regulatory T cell
TNF-α: Tumor necrosis factor α
IFN-γ: Interferon γ

## Acknowledgements

The authors thank Florin Tuluc and Jennifer Murry from the CHOP Flow Cytometry Core Laboratory and Fang Chen and Natalka Kengle from the Center for Immunotherapies at the University of Pennsylvania for their contributions to this study.

